# Edge Artificial Intelligence for real-time automatic quantification of filariasis in mobile microscopy

**DOI:** 10.1101/2023.08.02.23293538

**Authors:** Lin Lin, Elena Dacal, Nuria Díez, Claudia Carmona, Alexandra Martin Ramirez, Lourdes Barón Argos, David Bermejo-Peláez, Carla Caballero, Daniel Cuadrado, Oscar Darias, Jaime García-Villena, Alexander Bakardjiev, Maria Postigo, Ethan Recalde-Jaramillo, Maria Flores-Chavez, Andrés Santos, María Jesús Ledesma-Carbayo, José M. Rubio, Miguel Luengo-Oroz

## Abstract

Filariasis, a neglected tropical disease caused by roundworms, is a significant public health concern in many tropical countries. Microscopic examination of blood samples can detect and differentiate parasite species, but it is time consuming and requires expert microscopists, a resource that is not always available. In this context, artificial intelligence (AI) can assist in the diagnosis of this disease by automatically detecting and differentiating microfilarias. In line with the target product profile for lymphatic filariasis as defined by the World Health Organization, we developed an edge AI system running on a smartphone whose camera is aligned with the ocular of an optical microscope that detects and differentiates filarias species in real time without the internet connection. Our object detection algorithm that uses the Single-Shot Detection (SSD) MobileNet V2 detection model was developed with 115 cases, 85 cases with 1903 fields of view and 3342 labels for model training, and 30 cases with 484 fields of view and 873 labels for model validation before clinical validation, is able to detect microfilarias at 10x magnification and distinguishes four species of them at 40x magnification: *Loa loa, Mansonella perstans, Wuchereria bancrofti*, and *Brugia malayi*. We validated our augmented microscopy system in the clinical environment by replicating the diagnostic workflow encompassed examinations at 10x and 40x with the assistance of the AI models analyzing 18 samples with the AI running on a middle range smartphone. It achieved an overall precision of 94.14%, recall of 91.90% and F1 score of 93.01% for the screening algorithm and 95.46%, 97.81% and 96.62% for the species differentiation algorithm respectively. This innovative solution has the potential to support filariasis diagnosis and monitoring, particularly in resource-limited settings where access to expert technicians and laboratory equipment is scarce.

## 1. Introduction

Filariasis is a tropical infectious disease caused by roundworms (Phylum Nematoda). There are at least eight filarial worms that are hosted in humans. These are the causative agents of four types of diseases: lymphatic filariasis, which is caused by *Wuchereria bancrofti, Brugia malayi*, and *Brugia timori*; Onchocerciasis, caused by *Onchocerca volvulus*; loiasis, caused by *Loa loa*; and mansonellosis, caused by *Mansonella perstans, Mansonella ozzardi*, and *Mansonella streptocerca*. Among these, lymphatic filariasis and onchocerciasis have significant clinical and public health implications, while loiasis and *Mansonella perstans* have historically received less attention and are considered neglected diseases (1–3).

In 2000, the World Health Organization (WHO) launched the Global Programme for the Elimination of Lymphatic Filariasis (GPELF), which set the goal of eliminating lymphatic filariasis as a public health problem in 58 countries by 2030 (4). The program achieved a considerable reduction, but there are still 863 million people in 50 countries who require preventive chemotherapy (PC) (5). Similarly, onchocerciasis affects over 20.9 million people, with at least 220 million in need of PC (6). However, *L. loa* infection is hindering the elimination of lymphatic filariasis and onchocerciasis, as these diseases use ivermectin in massive drug administration (MDA), but ivermectin causes severe adverse effects when the individual has elevated levels of *L. loa* in the blood (7).

Studies have reported that *M. perstans* is the most prevalent filariasis in Africa, with more than 100 million people estimated to be infected and 600 million living in 33 high-risk countries (8), yet it is one of the most neglected filariasis (2,3,9), and there are no control programs for it.

The correct diagnosis and appropriate treatment are paramount for the effective control and elimination of parasites and their approach depends on the filarias type. WHO recommends utilizing the Alere Filariasis Test Strip (FTS) for all areas endemic for *W. bancrofti* and Brugia Rapid Test for all areas endemic for *Brugia spp*. However, these tests are species-specific and do not account for co-infections (10). Molecular diagnosis methods have also been applied in surveillance studies with good results but without possibilities to perform on site (11). Microscopy remains the most widely used technique for all filarial species, enabling the detection of microfilariae through blood smears or skin snips. The routine examination is the screening at low magnification (10x magnification) and then uses higher magnification to identify the species (e.g., 40x). The sample should be scanned completely at 10x magnification to report the sample as negative (12). Nonetheless, the diagnosis by microscopy is time-consuming and requires experienced microscopists, whose availability is not always assured (13,14). In that sense, different studies revealed the importance of mobile health (mHealth) to bring diagnostics to the point of care and scale access in low and middle-income countries (LMICs) (15–17). Notably, several investigations reported the use of mobile microscopy for parasite detection, such as LoaScope, which is a point-of-care microscope that detects *L. loa* microfilaria in blood smears automatically in video (18,19) or SchistoScope, a mobile phone microscope for the screening of *Schistosoma haematobium* (20).

A possible tool to address the lack of trained specialists is the detection of parasites in microscopic images using Artificial Intelligence (AI). AI is revolutionizing the medical field and can be applied in different medical subfields (21,22). The development of AI algorithms for microscopy depends on the digitization samples, which can be done using digital microscopes that have embedded cameras, converting a conventional optical microscope to a digital microscope using mobile phones or other image acquisition modules.

Numerous studies reported the detection of parasites in microscopical images, revealing the potential of AI in this task. Quinn *et al*. created one of the first deep learning algorithms for malaria image classification, for that, they used a 3D printed adapter that aligns the mobile phone camera to microscope eyepiece (23). Davidson *et al*. presented a study that detects Plasmodium-infected cells and classifies the stage obtaining a 98.5% average precision in detecting RBCs and 99.8% in classifying the detected cells into infected or uninfected. Images in this study were acquired by manually aligning the mobile phone camera to the microscope eyepiece (24). Similarly, Holmström *et al*. presented a deep learning algorithm for the detection of soil-transmitted helminths and *Schistosoma haematobium* with a custom microscope scanner (25). Dacal *et al*. presented an object detection algorithm for STH that runs on a smartphone (27). Oyibo *et al*. presented an automated microscope with AI for the detection of *Schistosoma haematobium* (28). Dedhiya *et al*. introduced the first study that uses machine learning over thermal imaging to predict the viability of onchocerca worms (29). D’Ambrosio *et al*. presented an algorithm which detects *L. loa* microfilaria in video, without distinguishing species. They correlated the automated counts with manual counts and achieved 94% specificity and 100% sensitivity (18). Elvana *et al*. presented a lymphatic filariasis detection system using image analysis, achieving an accuracy of 70% (30). As far as we know, there have been very few attempts to deploy edge-AI systems using deep learning which are able to support in real-time and without connectivity the analysis of optical microscopy images for filaria detection with species differentiation and more broadly NTDs diagnostics.

The objective of this study is to propose, develop, and pilot a system for real-time, automatic detection and quantification of filariasis using an edge AI model. The proposed system aims to assist the screening and species differentiation of four worm species (*L. loa, M. perstans, W. bancrofti* and *B. malayi*) in blood smears for filariasis. For that, we proposed a pipeline with the following modules: the digitization of smear samples with smartphones coupled to a microscope through a 3D-printed device; sample analysis and data labeling in a telemedicine platform for training of an AI algorithm; integration of the trained algorithm on the smartphone to assist the diagnosis and validation of the model in a clinical environment.

## 2. Materials and Methods

### 2.1 Ethics statement

Ethical approval was obtained from the Research Ethics Committee (REC) Instituto de Salud Carlos III, Spain (CEI PI 74_2020).

### 2.2 Overview of the methodology

The study was conducted in two distinct phases. The initial phase involved digitizing blood smear samples to construct the database for the development of AI algorithms with 115 samples. In the subsequent phase, the AI model was integrated on the smartphone and a pilot study was conducted to evaluate the AI’s performance on real world settings with a new dataset of 18 samples.

All preparations included in the study were appropriately stained, positive and with well-preserved parasite morphology. Different levels of parasitemia ensured the possibility of collecting positive and negative fields of view. In addition to the image, other information regarding each preparation is also collected, such as the result obtained through alternative methods like polymerase chain reaction (PCR) and/or conventional microscopy, the staining type and where it was a thin spread or a thick drop.

### 2.3 Creating a Filariasis differentiation AI model

#### 2.3.1 Digitalize samples

In the initial phase, a total of 115 sample smears from 115 different subjects were collected from the sample collection of the Malaria and Emerging Protozoa Unit of the Instituto de Salud Carlos III (Spain). In addition, all preparations have been previously anonymized without the possibility of reverse coding. 112 of them were stained with Giemsa and 3 of them with Panopticon. Case distributions were presented in **Table 1**.

**Table 1:**
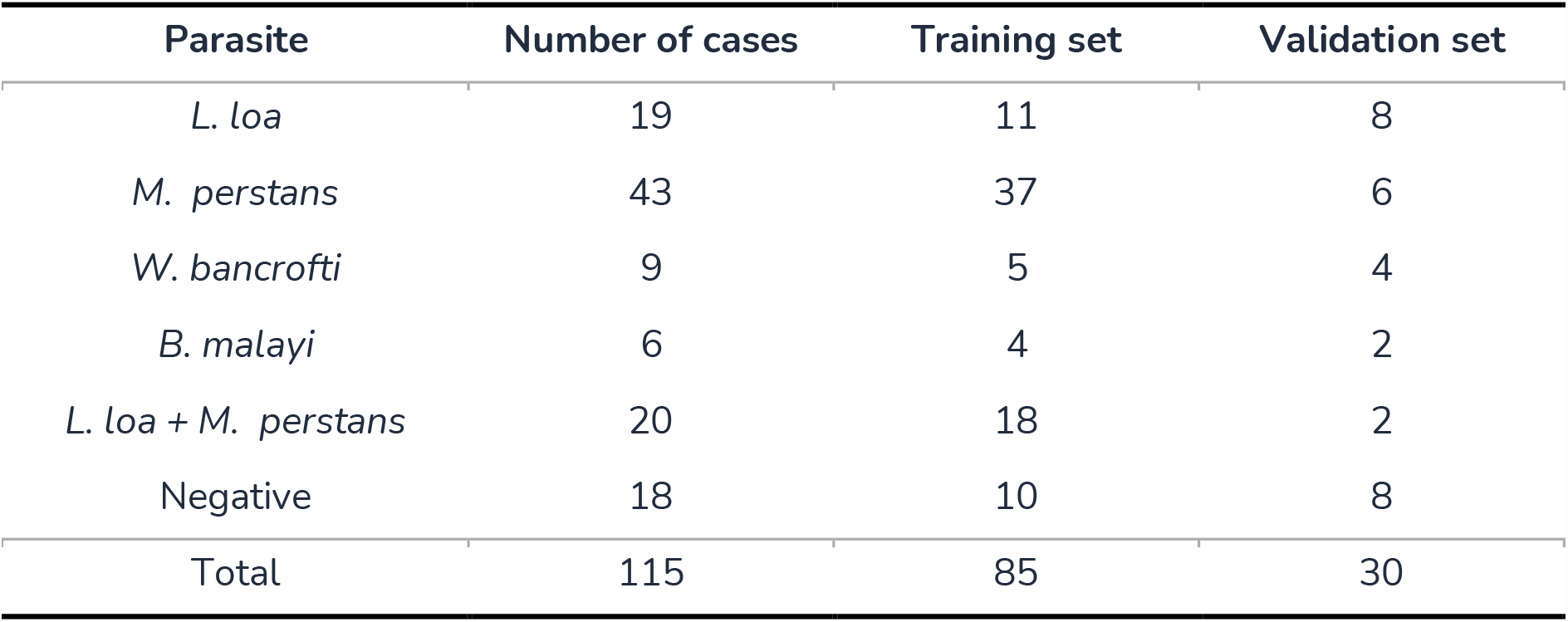
Cases included in the first phase and the training-validation split.

Images were digitized simulating the real diagnostic workflow, with a system previously described in Dacal *et al*. (27). Briefly, this system uses a 3D printed device that allows coupling a mobile phone with a conventional optical microscope by aligning the smartphone camera with the objective of the microscope to acquire images, and that converts any conventional microscope into a digital microscope. Following the conventional workflow, the analyst scanned the samples at 10x magnification and captured photos of fields containing structures compatible with filarial parasites. Subsequently, the objective was switched to 40x magnification, and photos of each detected parasite were taken. Slides were digitized using four different smartphone models (Huawei Ascend G7; (n=95 cases), Redmi Note7 (n=13 cases), Samsung Galaxy A32; (n= 5 cases), LG X Power K220; (n=1 case), Huawei Nova 5T (n=1 case)). 873 images of 10x and 1514 images of 40x were captured.

To ensure complete separation between the training set and validation set, we implemented a case-level split for the digitized cases. This approach effectively prevents the inclusion of validation set data in the training set, thus addressing the issue of overfitted performance, and ensuring accurate reporting.

#### 2.3.2 Labeling data

All acquired images were transferred from the smartphone to a telemedicine platform via mobile network, so that the images are stored and presented in an easy-to-use dashboard that allows their visualization, management, and labeling (**Figure 1**). In this web platform, standard clinical and analysis protocols were translated into digital tasks that were adapted to the clinical case and disease under study.

**Figure 1:**
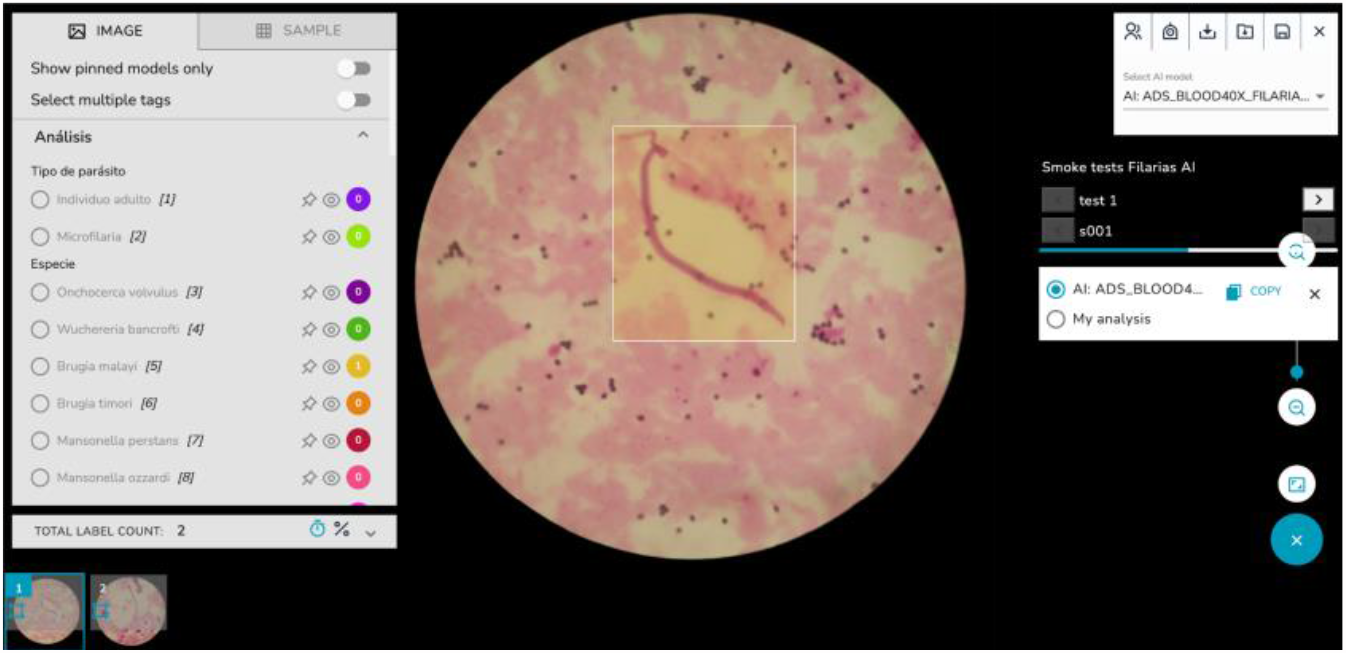
The telemedicine platform facilitates image visualization, management, and labeling. When an AI algorithm is deployed, analysts have the option to review the predictions rather than starting the labeling process from scratch.

The annotation protocol was based on the placement of bounding boxes around the identified parasites. All visible parasites in the image were labeled by two analysts and reviewed by an expert. At 10x magnification, as the species can’t be identified, all detected parasites belong to the microfilariae class. A total of 2293 parasites were located from 837 images. At 40x magnification, the parasite species were annotated with their corresponding class. A total of 1651 parasites were tagged from 1514 images. In addition, some artifacts that have a similar appearance to the parasite were labeled, which serves as a hard negative for the algorithm training.

The labeled data was divided into a training set for model development and a validation set for selecting the best model, as shown in **Table 2**. The training set for 10x images consists of 1965 microfilarias from 700 images, while the validation set for 10x images contains 328 microfilarias from 173 images. In the training set for 40x images, there are 906 *L. loa*, 378 *M. perstans*, 35 *W. bancrofti*, and 58 *B. malayi* parasites from 1203 images, while the validation set includes 38 *L. loa*, 102 *M. perstans*, 29 *W. bancrofti*, and 5 *B. malayi* parasites from 311 images.

**Table 2.** Label distribution of microfilaria species in the training and validation sets.

| Labels | total | training | validation |
| --- | --- | --- | --- |
| Microfilaria | 2293 | 1965 | 328 |
| *L. loa* | 1044 | 906 | 138 |
| *M. perstans* | 480 | 378 | 102 |
| *W. bancrofti* | 64 | 35 | 29 |
| *B. malayi* | 63 | 58 | 5 |

#### 2.3.3 Creating the AI Model

A requirement for our AI model is that it can work offline or in limited bandwidth settings. To fulfill this requirement, we selected a lightweight model that can be run on a smartphone in real-time without internet connection. Given the multifaceted nature of the task, encompassing object localization, classification and counting, an object detection algorithm would be an appropriate solution. Specifically, we employed the Single-Shot Detection (SSD) MobileNet V2 detection model with a feature pyramid network as feature extractor, shared box prediction and focal loss (31–33). Tensorflow object detection application programming interface (API) was used for model training because tensorflow has natively optimized the model to be executed in mobile phones and edge devices (34). Given the relatively small size of our dataset, we used a pre-trained model that was trained with the COCO image database (35) and fine-tuned for this use case.

Two distinct algorithms were developed. The first algorithm, designed for screening at 10x magnification, focuses solely on detecting the presence of microfilariae. The second algorithm, developed for microfilaria species differentiation at 40x magnification, aims to classify the detected microfilariae into four species: *L. loa, M. perstans, W. bancrofti, and B. malayi*.

Given the alignment of the smartphone with the microscope eyepiece, the area visualized by the mobile phone is limited to a circular area, as depicted in **Figure 2**. In order to exclude non-informative regions (e.g., black areas), and to present other relevant information on the mobile phone screen (e.g., label count, AI activation, etc), we decided to use square images instead of rectangular images.

**Figure 2.**
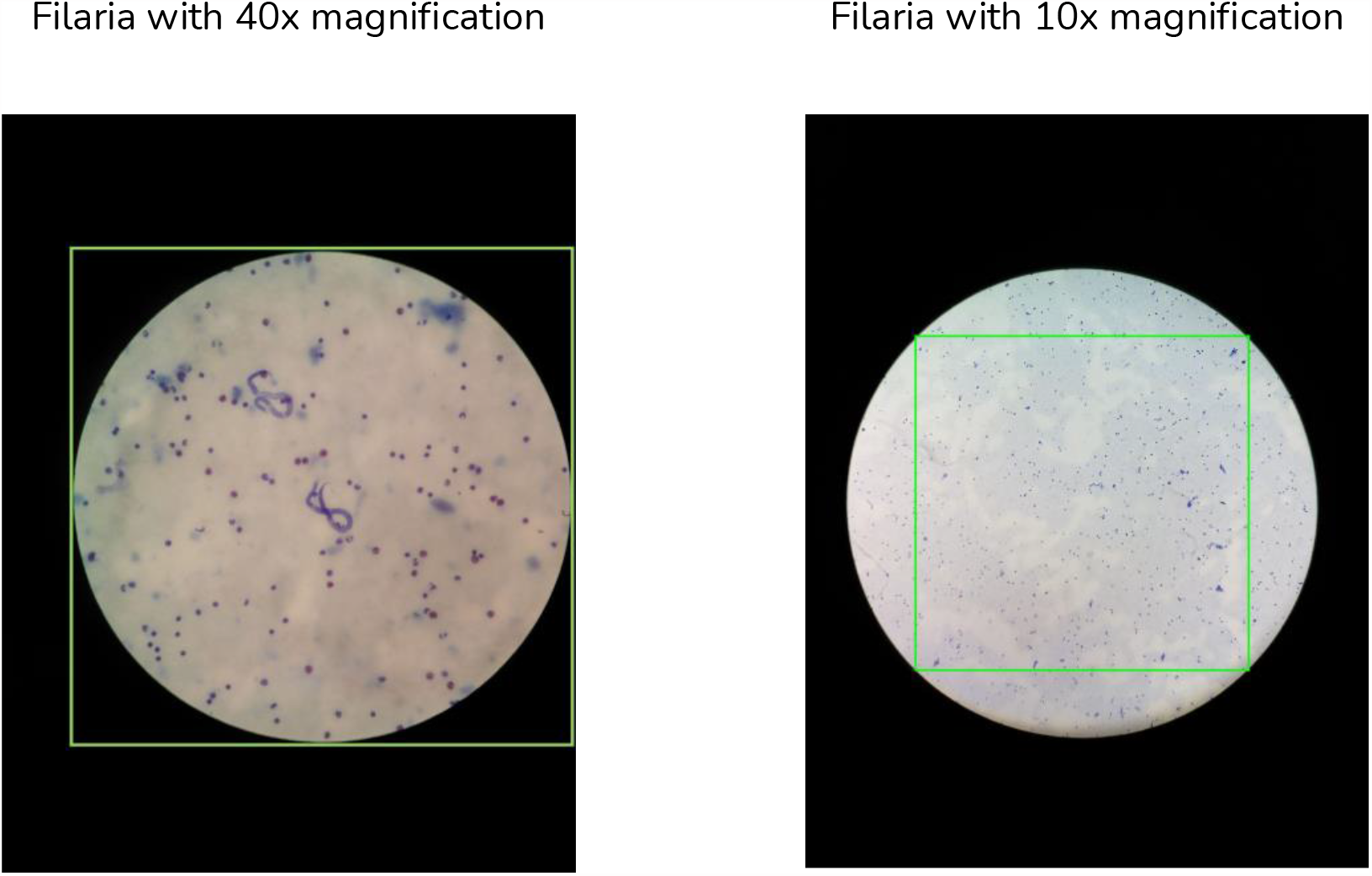
Left): example input of the species differentiation algorithm. Right): green rectangle represents the sliding windows size

For the species differentiation algorithm that works with 40x magnification, we initially identify the circular region within the image and extract a square image encompassing the entire field of view, as illustrated on **Figure 2 left**. Subsequently, the cropped region was resized to 640×640 pixels. The reviewed data was splitted into two sets at case level as described above.

To address the imbalance nature of the dataset, oversampling of the minority classes was employed by generating mosaic images, which consists of cropping a 320×320 pixel patches that contains at least one parasite, and blending 4 images to create a new image of 640×640 pixels. After augmentation, the training set contains 1116 *L. loa*, 378 *M. perstans*, 480 *W. bancrofti* and 533 *B. malayi*. Additional image augmentation, including random flip, rotation, crop, and brightness, hue and saturation adjustment, was applied during training to enhance the model’s robustness.

In the screening algorithm that works with 10x magnification, a distinct strategy was implemented for image cropping in comparison to the species differentiation algorithm. Given the relatively small size of the parasite in 10x magnification, its visualization and detection pose challenges for both human analysts and AI systems, necessitating the use of zoom. To optimize the visibility of the parasite and maximize the size of the image, we decided to crop the original image to the square inscribed within the circle as depicted in **Figure 2 right**. As it can be appreciated, a single crop of the inner square leaves some valuable information out. To overcome this limitation, we employed the sliding window technique, where 4 patches were generated for each image, ensuring that all the information within the field of view is represented. Then, patches were resized to 640×640 to fit the requirements set by the network used. The same data augmentation was applied as in the species differentiation algorithm. After data augmentation, the number of microfilarias in the training set increased from 328 to 10847, whereas the validation set was unmodified.

### 2.4 Validation of the AI model in a clinical setting

To assess the usability and performance of the proposed system within the clinical workflow, a field validation was conducted. Initially, the AI model was deployed on the smartphone, followed by an evaluation of the model’s performance using an analysis workflow with 18 samples that were not used for training nor validation of the model.

#### 2.4.1 Deployment and integration of technology

The AI mobile model was optimized using post-training quantization, which is a conversion technique that reduces model size while also improving CPU and hardware accelerator latency, with little degradation on model accuracy. Subsequently, the model was converted into the Tensorflow Lite format. The model then is embedded into the custom application to perform the image analysis in real time. Speed test is conducted on two smartphones: the BQ Aquaris X2 and the Samsung S9. The execution time on BQ Aquaris X2 on CPU is 1400 milliseconds for a single image, while the Samsung S9, utilizing the GPU, accomplished the task in 610 milliseconds.

To facilitate the process of digitization and AI-assisted analysis, a customized Android application was developed. This application records both clinical data and images. While the user visualizes the image on the mobile phone screen, the selected AI algorithm, screening or species differentiation is running in real time depending on the magnification used (10x or 40x), generating predictions for the corresponding frame, and outlining the detected parasites within bounding boxes. When the user takes a photo, both the images and the prediction are saved, and the parasite detection counter is incremented no matter if the prediction is correct or not. In the case that the user finds parasites not detected by the AI algorithm, they can tap on the button of the corresponding label to increase the count of this parasite. Once the analysis is finished, this information is uploaded to the telemedicine platform, allowing users to review and correct the prediction and share information. **Figure 3** explains how the smartphone is attached to the conventional and the screening and species differentiation algorithm running on the smartphone. In addition, the **Video 1** on supplementary data showcases the whole AI system including hardware. As the video demonstrates, the smartphone camera is aligned to the microscope eyepiece with the assistance of an adapter, with AI running in real time, the analyst moves the sample and analyzes it with AI assistance. **Video 2** in the supplementary data provides a screen recording that offers a more detailed perspective of AI detecting microfilarias at 10x magnification and differentiating species at 40x magnification.

**Figure 3:**
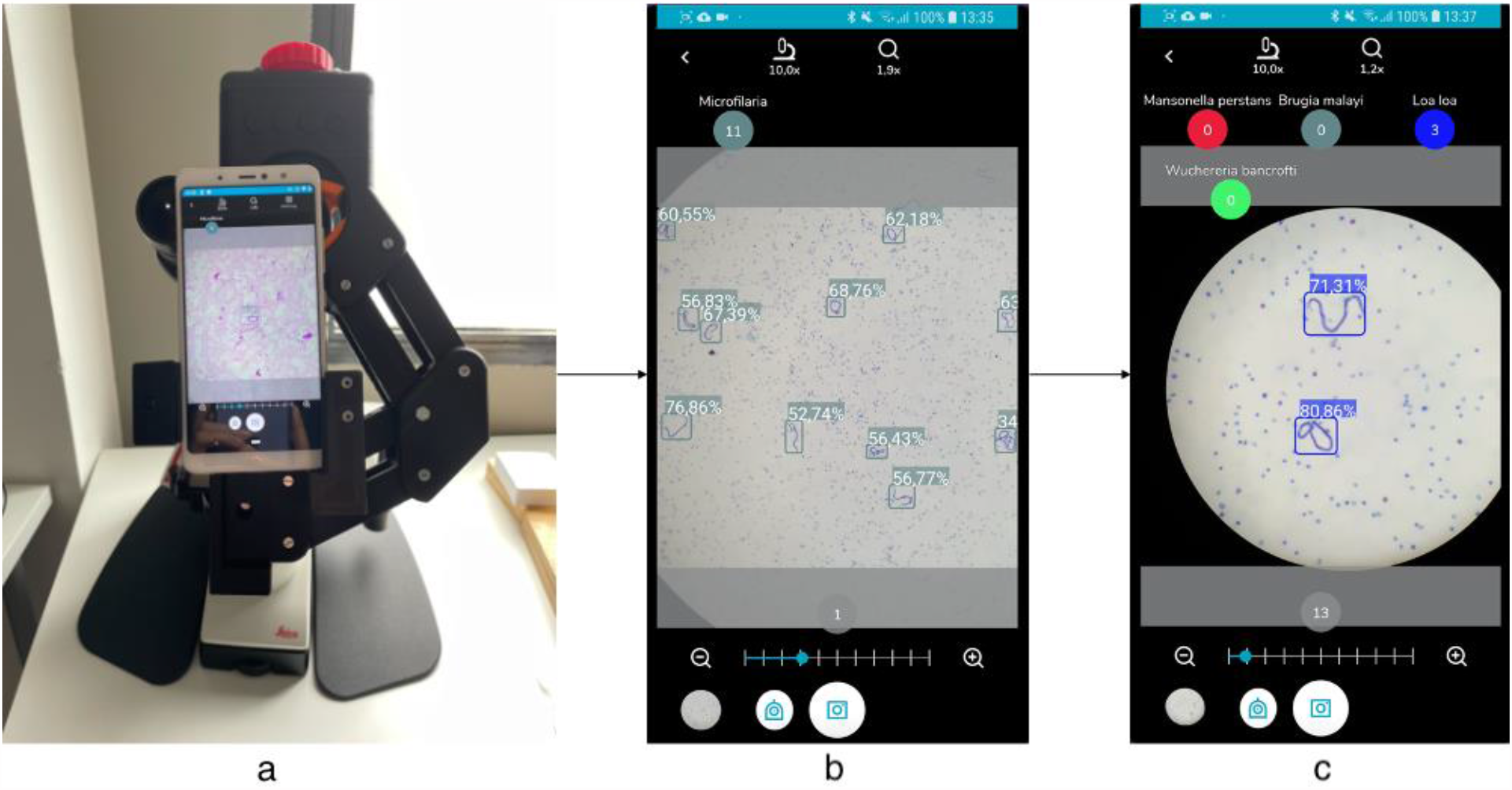
(a) smartphone attached to the conventional microscope with a 3D printed adapter. (b) screening algorithm working on the smartphone. (c) species differentiation working on the smartphone.

#### 2.4.2 Experiment-Pilot replicating diagnostic workflow

To assess the performance of the AI models, a real-time pilot study was conducted to evaluate the effectiveness of the edge AI system in assisting parasite detection through the mobile application.

To pilot the proposed system, the algorithm developed in the first phase was integrated on the mobile phone and the telemedicine platform to be validated. For all selected samples (N=18), **Figure 4** represents the ideal workflow: with the AI algorithm operating in real time, the analyst examines the complete sample using a 10x magnification objective. Depending on whether the algorithm detects a parasite, different actions are taken. When a parasite is identified by the AI algorithm, the analyst captures a photo and the detected parasites are automatically counted by the app, and switches to a 40x magnification objective. At this point, the species differentiation algorithm is activated to discern the specific species of the detected parasites, a photo is taken to count the parasite. In cases where parasites are present on the screen but not detected by the screening AI, the analyst manually adds the count by tapping on the corresponding label. Since the mobile application did not allow modification of the incorrect prediction, both images and the mobile prediction were uploaded to the telemedicine platform for further correction and validation. The results were independently reviewed by two analysts: analyst A, a junior researcher in parasitology, who analyzed images on real time using the mobile application; and analyst B, an expert in microscopy of infectious diseases and who only reviewed the digitized image on the telemedicine platform.

**Figure 4.**
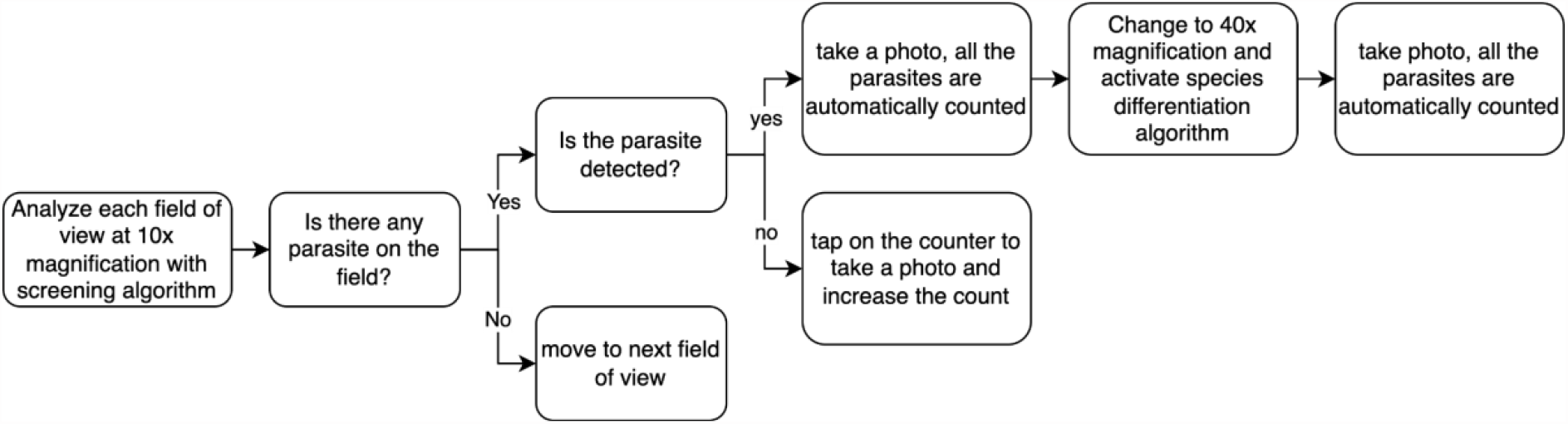
Schema represents the validation workflow of AI assisted filariae detection. At least 3 images of negative fields were acquired for each sample.

The evaluation of the algorithm’s performance was based on precision (P), which measures the proportion of correctly identified objects among all the objects predicted by the model; recall (R), which measures the proportion of the correctly identified objects among all the ground truth objects; and F1 score, a combined metric that takes into account both precision and recall to provide a single value that represents the overall performance. Object detection algorithms have capabilities that go beyond classification algorithms, being able to detect multiple objects as well as their location and size within the image, in the form of bounding boxes. Therefore, to compute those metrics, additional considerations must be put in place. For each proposed bounding box with confidence score greater than 50%, it is considered as a true positive (TP) if the intersection over union with the ground truth is greater than 0.5 and the class is correct. Conversely, if the predicted area corresponds to artifacts or other parasites class then it is considered as false positive (FP). Furthermore, ground truth boxes that were not proposed by the algorithm were categorized as false negatives (FN). True negative (TN) were not computed as all areas without predictions are considered TN.

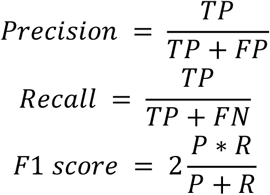

## 3. Results

### 3.1 Evaluation of the AI model performance

The performance assessment of the model was conducted on the validation set with 30 cases as described in **Table 1**. The screening algorithm, designed to work with 10x magnification, achieved a precision of 88.17%, recall of 91.62%, and an F1 score of 89.85%. On the other hand, the species differentiation algorithm achieved a weighted precision of 84.08%, recall of 95.33%, and an F1 score of 94.70%. Breaking down the results per class, the precision rates were 94.85% for *L. loa*, 97.03% for *M. perstans*, 94.00% for *W. bancrofti*, and 66.67% for *B. malayi*. The corresponding recall rates were 93.48%, 96.08%, 97.92%, and 92.31% respectively.

The resulting confusion matrix of the species differentiation algorithm that works with 40x magnification is presented in **Table 3**. It should be noted that the AI algorithm was not specifically trained with artifact labels, which are areas on the image that may look like a parasite (e.g., mycelium, fibers), but the model may occasionally predict the artifact region as a parasite (false positive) where the analyst did not assign a label.

**Table 3:** Confusion matrix of the species differentiation algorithm (40x) on the validation set, each row represents the ground truth, and each column represents the prediction. The model may predict the artifacts as a parasite (false positive), but the analysts did not label all artifacts.

|  |  | AI Predictions |  |  |  |  |
| --- | --- | --- | --- | --- | --- | --- |
|  |  | Artifact | *L. loa* | *M. perstans* | *W. bancrofti* | *B. malayi* |
| Ground Truth | Artifact | - | 7 | 3 | 2 | 3 |
|  | *L. loa* | 6 | 129 | 0 | 1 | 1 |
|  | *M. perstans* | 4 | 0 | 98 | 0 | 0 |
|  | *W. bancrofti* | 0 | 0 | 0 | 47 | 1 |
|  | *B. malayi* | 1 | 0 | 0 | 0 | 12 |

### 3.2 Validation of the AI-assisted mobile app

For the pilot study, a total of independent 18 samples from different subjects with respect to the ones used for training and validation were analyzed with AI assistance on the mobile phone by analyst A. 452 field of views of 10x magnification and 624 field of views of 40x magnification were analyzed on the mobile phone, uploaded to the telemedicine platform, and reviewed by another analyst, generating the ground truth to evaluate the model performance in real time.

To assess the potential benefits of reviewing AI analysis and its impact on inter-observer variability and time, we shuffled and split the uploaded images into four groups, 10x magnification with AI and without AI assistance (232 images with AI and 220 images without AI), and 40x magnification with and without AI assistance (320 images with AI and 304 images without AI). Both analysts analyze the two groups without AI assistance -10x without AI and 40x without AI - from scratch and analyze the two groups with AI assistance -10x with AI and 40x with AI - by reviewing the prediction generated by the AI on the smartphone. This allowed us to investigate the potential time-saving benefits and the potential reduction in inter-observer variability provided by AI assistance.

#### 3.2.1 Real-time AI-Performance

The analysis conducted by analyst B, who has greater expertise compared to analyst A, was considered as the ground truth for our evaluation. According to analyst B, at 10x magnification, out of the 452 images examined, 280 were identified as positive, indicating the presence of at least one parasite, while 172 were determined to be negative. The parasite count reported by analyst B and the AI for each image is significantly correlated, with a pearson correlation coefficient of 0.984. At parasite level, the screening algorithm achieved an overall performance of 94.14% precision, 91.90% recall, and 93.01% F1-score. In the context of differentiating between parasite species, according to analyst B, out of the 624 images assessed, 511 were classified as positive. The parasite count reported by analyst B and the AI for each image is also significantly correlated for species differentiation algorithm, with a pearson correlation coefficient of 0.953. The AI algorithm demonstrated an overall precision of 95.46%, recall of 97.81%, and F1-score of 96.62% in this regard. The per-class precision values were determined as 98.80% for *L. loa*, 60.00% for *M. perstans*, 100.00% for *W. bancrofti*, and 58.97% for *B. malayi*. The corresponding recall rates were calculated as 98.50%, 100.00%, 76.00%, and 100.00%, respectively. **Table 4** presents the confusion matrix of the AI model in relation to analyst B’s analysis.

**Table 4:**
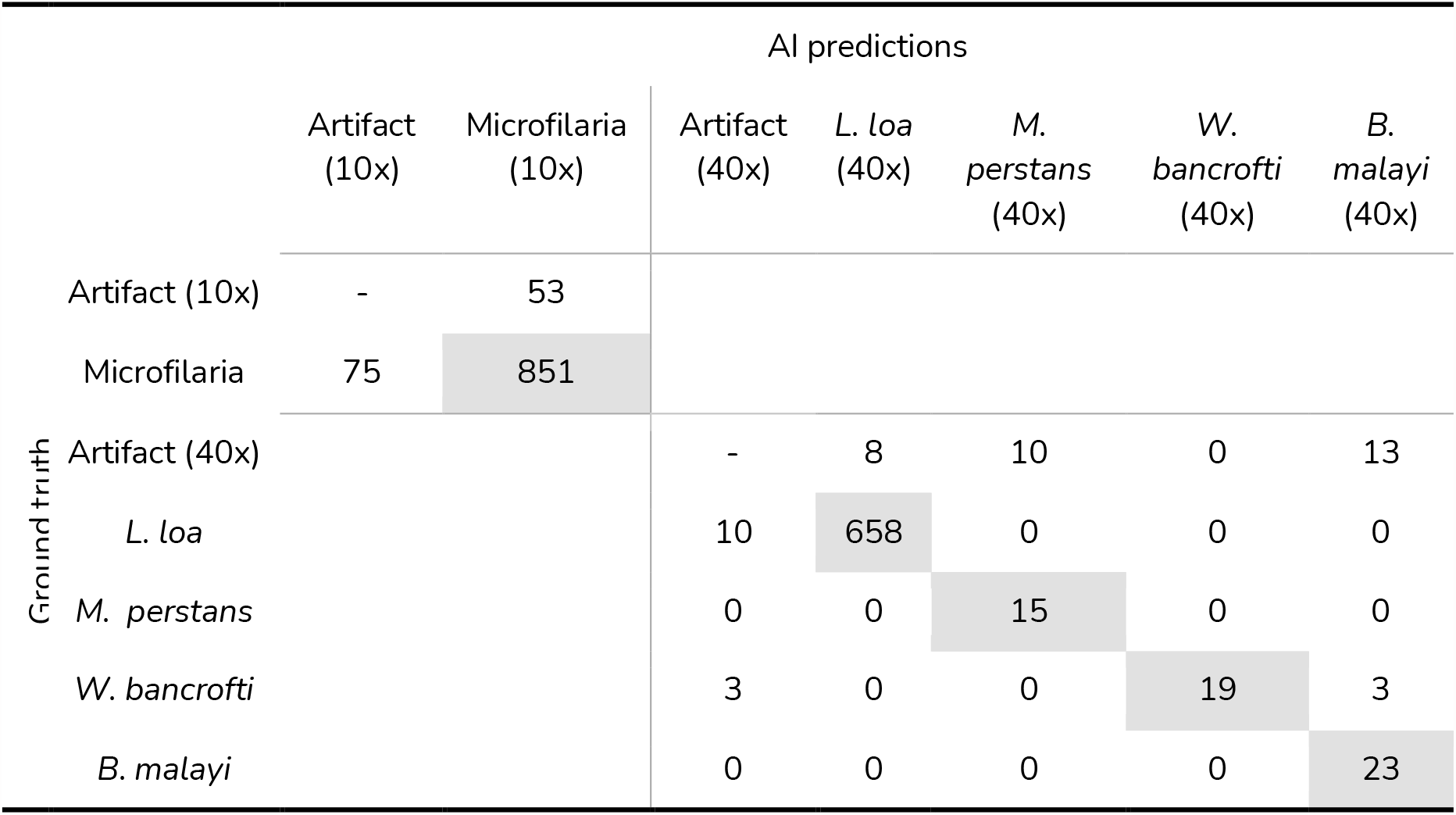
Performance of the AI algorithm on pilot study using mobile phone. Each row represents the ground truth and each column represents the AI prediction.

**Table 5:** Inter-observer agreement of detected parasites when analyzing with and without AI assistance of 2 experts and of the AI. * indicates that the p-value is smaller than 0.001.

|  | With AI | Without AI |
| --- | --- | --- |
| 10x | 0.994* | 0.990* |
| 40x | 0.997* | 0.992* |

**Table 6:** Average analysis time in seconds for both analysts with and without AI assistance for each image.

|  | without AI assistance | with AI assistance |
| --- | --- | --- |
| Analyst A | 23.5 | 3.5 |
| Analyst B | 12 | 12 |

#### 3.2.2 Inter-observer variability

To assess inter-observer variability, we compared the total number of parasites detected in each sample by analyst B and analyst A, both with and without AI assistance. Additionally, we compared the parasite count of each image generated by the AI system with the count provided by analyst B (considered as the ground truth). This comparison allowed us to analyze the performance and agreement between the analysts and the AI system in identifying and quantifying parasites.

The results reported by both analysts are strongly correlated. The Pearson correlation coefficient between analyst A and analyst B when analyzing without AI is 0.989 for the screening algorithm and 0.992 for species differentiation algorithm. Similarly, when analyzing with AI assistance, the correlation coefficients are 0.994 and 0.997 respectively. Notably, the correlation coefficients are slightly higher when analysts utilize AI assistance during their assessments.

Furthermore, it is noteworthy that there is a high correlation between the parasite counts reported by the AI model and analyst B. The minimum Pearson correlation coefficient observed in this comparison was 0.928, further indicating a strong correlation between their reported counts.

#### 3.2.3 Analysis time

In addition to evaluating the performance of the edge AI system, we also analyzed the potential time-saving effect of AI assistance on the telemedicine platform. We compared the time required for analysts to review AI predictions versus the time needed for labeling from scratch. Both analysts were asked to review 524 images without AI assistance, and then 524 different images with AI assistance. For Analyst A, the analysis time significantly decreased from an average of 23.5 seconds per image to just 3.5 seconds per image when utilizing AI assistance. However, it should be noted that the analysis time for Analyst B remained unchanged.

## Discussion and Conclusion

This study introduces the first real-time edge AI deployment on smartphones to assist in the screening and species differentiation for filarial samples in mobile microscopy and validated it in a clinical setting. To create and validate the AI powered mobile application, we proposed a methodology that encompasses an image digitization system, a telemedicine platform to visualize and annotate images, a training and deployment pipeline, and an Android application to deploy AI models.

Diagnosis is an essential part of the monitorization of the effect of MDA, which is a recommended strategy to control or eliminate several neglected tropical diseases, including filariasis. Microscopy is a widely used technique for filariasis diagnosis, as it can distinguish parasite species, but it requires expert microscopists, and is time-consuming. Very few studies attempted to automatize filarias parasite detection detecting microfilarias, without distinguishing species (18,30). With respect to these prior works, our proposal allows us to replicate the full diagnostic workflow including 10x and 40x examinations, successfully distinguishing between different microfilariae species, making it particularly valuable in co-endemic areas where multiple species are prevalent. Our system also operates in real-time without the internet connection, enabling its deployment at the point of care and not relying on expensive or hard to find hardware as it can be utilized with any conventional microscope and low-to middle-end mobile phones, making it accessible and affordable. The system is easily scalable, as it is deployed on smartphones.

The AI system that we propose follows the conventional workflow, screening the sample at 10x magnification and differentiating species at 40x magnification. Hence, two algorithms were deployed for each use case using 85 samples, which were first validated on 30 samples to assess the model performance and then deployed to the clinical environment to evaluate the whole system usability. The validation in the clinical environment was conducted by analyzing 18 samples with the AI model running on mobile phone in real time, achieving an overall precision of 94.14%, recall of 91.90% and F1 score of 93.01% for the screening algorithm and 95.46%, 97.81% and 96.62% for the species differentiation algorithm respectively.

In the inter-observer variability and analysis time comparison, we found that with AI assistance the correlation between two analysts increased slightly, and the analysis time reduced for the junior researcher in parasitology while it remained unchanged for the expert in infectious diseases microscopy.

It is important to acknowledge that our study has a limited sample size in general, especially for *W. bancrofti*, and *B. malayi* (35, 58 labels for training respectively). Despite that, our algorithm achieved high precision and recall, even though the performance fluctuates a lot for minority classes. Additionally, the fact that all samples come from one research center may introduce bias and reduce generalizability of our algorithm, performing worse in samples from other centers, due to the sample preparation, etc. To address these limitations, future research should include multi-centric study, including training and validating on samples from different research centers and involving more analysts. Such an extensive validation process would help to assess the robustness and generalizability of the AI system across various real-world settings and conditions.

In conclusion, the presented system can assist the diagnosis of filariasis in resource-constrained settings, particularly when healthcare workers are scarce, by transforming any optical microscope into an intelligent point-of-care device. The system is easily scalable, as it is deployed on smartphones. This approach could reduce the dependency of highly specialized personnel as we can empower community health workers to contribute to filariasis control. Additionally, the system’s telemedicine platform provides the opportunity for seeking second opinions and quality control in cases of diagnostic uncertainty, enhancing overall accuracy. The platform also can be used as an epidemiological surveillance platform, contributing to the tracking and the monitoring of the prevalence and distribution of filariasis. Furthermore, our system can be expanded to other neglected tropical diseases by collecting samples of other diseases, with the vision of creating a universal AI model for parasite detection. We also believe that future AI supporting systems will be multi-modal, incorporating a wide range of clinical inputs from diverse data sources beyond imaging, such as medical text or speech, enhancing the accuracy and generating comprehensible diagnostic interfaces reports (36,37). The current AI revolution in medicine should also be viewed as an opportunity for NTDs.

## Data Availability Statement

Images and labels can be shared for research purposes upon request. Please contact Miguel Luengo-Oroz.

## Acknowledgements

This project has received funding from the European Union’s Horizon 2020 research and innovation programme under grant agreement No 881062. LL was supported by a predoctoral grant IND2019/TIC-17167 (Comunidad de Madrid). This work was supported by the Bill and Melinda Gates Foundation (grant number Edge-Spot project INV-051355).

## Author contributions

LL, ED, DB-P, MP, JMR, MJL-C, MLO conceived the concept of this work, LL, ED DB-P, ND CCar, AMR, LBA contributed to data acquisition and curation, C Car, AMR, LBA, DC OD, JG-V, ER-J, MFC, JMR provided study resources LL, ED, CCar, AMR, LBA, DB-P, MJL-C, JMR, MLO participated in the investigation, LL, ED, MJL-C, JMR, MLO designed and developed the methodology and created the models LL, DB-P, CCab, DC, OD, JG-V, AB, ER-J contributed to creating softwares that were used in this study, MLO, AS, MJL-C, JMR contributed as supervisors LL, ED, CCar, AMR, LBA, JMR, MLO contributed to the validation of the study LL and ED wrote the manuscript and all authors reviewed and edited the manuscript.

## Supplementary material

Video 1: [Demonstration of edge AI system for filariasis detection](https://drive.google.com/file/d/1XhlytiGwDQ4XeGKQaI7lKSvJXsm6y68A/view).

Video 2: [Screen recording showing automatic microfilarias detection at 10x and species differentiation at 40x magnification](https://drive.google.com/file/d/1dKM8RKYyFeK7rigC9PovDToP6tfOUkWO/view?usp=drive_link)

